# Development and optimization of a new method for direct extraction of SARS-CoV-2 RNA from municipal wastewater using magnetic beads

**DOI:** 10.1101/2020.12.04.20237230

**Authors:** Ana L. Parra Guardado, Crystal L. Sweeney, Emalie Hayes, Benjamin F. Trueman, Yannan Huang, Rob C. Jamieson, Jennie L. Rand, Graham A. Gagnon, Amina K. Stoddart

**Affiliations:** Centre for Water Resources Studies, Faculty of Engineering, Dalhousie University, Halifax, NS, Canada; Acadia University, Wolfville, NS, Canada

**Keywords:** SARS-CoV-2, RNA extraction, magnetic beads, wastewater surveillance, COVID-19, RT-qPCR

## Abstract

The use of magnetic beads in the extraction of nucleic acids from wastewater is presented as an approach to simplify extraction techniques for the detection of SARS-CoV-2 viral fragments in wastewater. In particular, this paper describes the development and optimization of a direct method for extracting SARS-CoV-2 RNA from municipal wastewater using magnetic beads. The recovery efficiency of the method using Accuplex SARS-CoV-2 Positive Reference Material (ASCV-2) was examined. Method factors assessed were sample volume, concentration of magnetic bead mix, elution temperature, and water matrix (deionized (DI) water and wastewater). The combination of optimized method parameters that resulted in the highest RNA recovery in both DI water (26.0 ± 0.8%) and wastewater (11.8 ± 1.4%) was a sample volume of 1.0 mL, a magnetic beads concentration of 100 µL mL^-1^ sample, and an elution temperature of 60 °C. The performance of this optimized method was further assessed in recovery experiments using wastewater samples spiked at 1.8×10^6^ and 1.8×10^4^ gene copies L^-1^ (GU L^-1^) with Gamma Inactivated SARS-COV-2 (GI-SCV-2) and 1.0×10^6^ and 1.0×10^4^ infectious units L^-1^ of Human Coronavirus 229E (HCV 229E) as viral surrogates. Recoveries of 86.1 and 4.6% were achieved for wastewater samples spiked with GI-SCV-2 at low and high concentrations, respectively. In assessing the effects of wastewater pre-filtration and addition of DL-Dithiothreitol (DTT, used to inactivate RNases that may degrade RNA) on recovery efficiency of ASCV-2, the magnetic bead-based extraction protocol performed optimally with unfiltered wastewater without DTT (recovery = 17.4 ± 0.4%). The method limit of detection (MLOD) for ASCV-2 recovered from pre-filtered wastewater was determined to be 4.6×10^4^ GU L^-1^ (95% degree of confidence). Using this optimized magnetic bead-based extraction protocol, the presence of SARS-CoV-2 RNA was verified in wastewater collected from sewershed locations in Atlantic Canada. This emerging RNA extraction method is direct, rapid, and does not require the use of specialized equipment, thus offering advantageous application for laboratories with limited resources. As such, this method is an indispensable tool in the monitoring of wastewater for SARS-CoV-2 to potentially understand COVID-19 infection occurrence within communities and inform public health leaders.

**Graphical Abstract:** 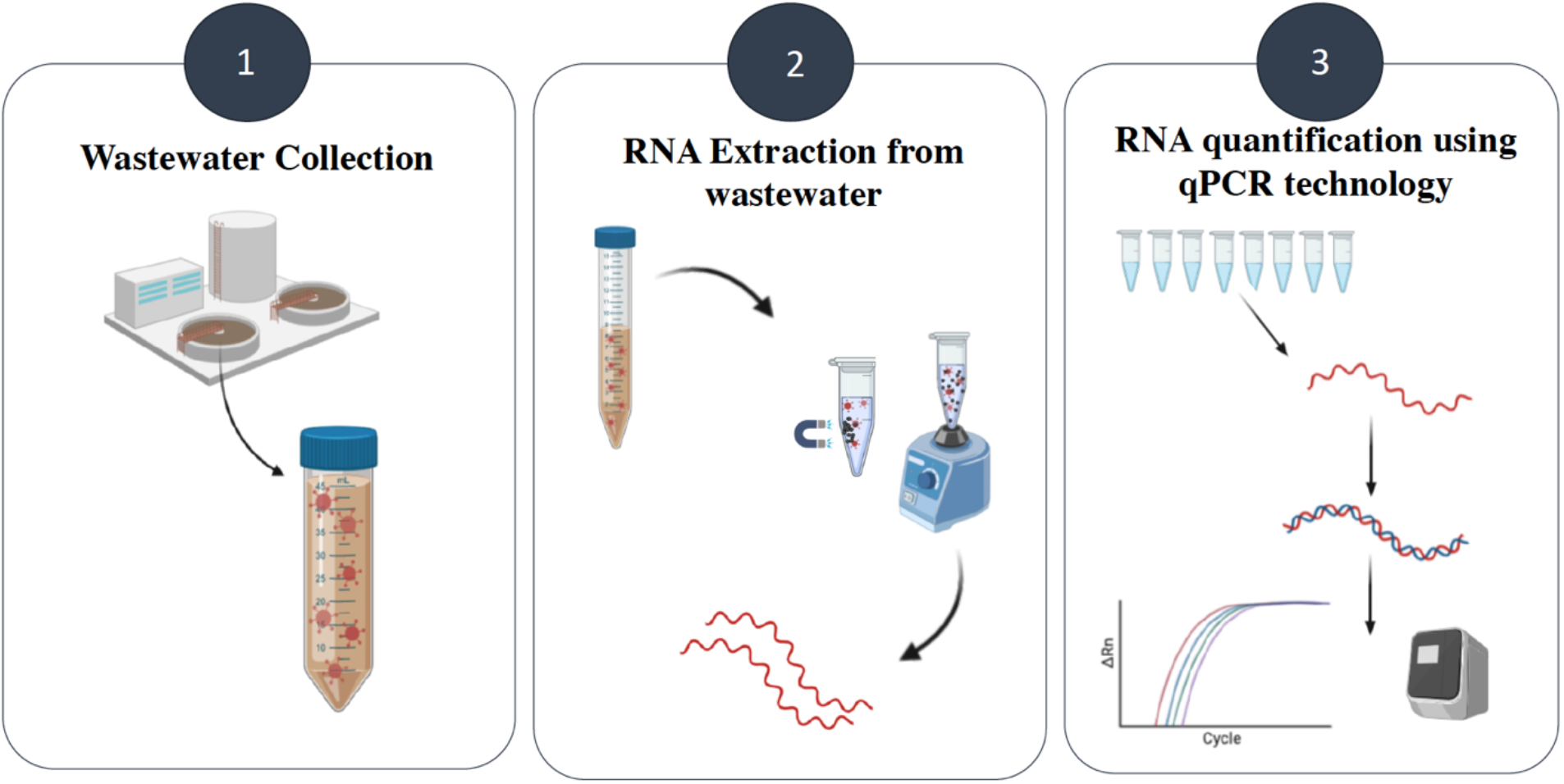

## 1. INTRODUCTION

The coronavirus disease 2019 (COVID-19) caused by the novel severe acute respiratory syndrome coronavirus 2 (SARS-CoV-2) continues to spread worldwide and has claimed the lives of over 1.3 million people as of 20 November 2020 (World Health Organization, 2020). SARS-CoV-2 is an enveloped, positively charged, single-stranded ribonucleic acid (RNA) virus (60 to 140 nm) of the beta coronavirus genus (Tian et al., 2020; Zhu et al., 2020). Although COVID-19 is characterized as a respiratory illness, many infected patients also present with gastrointestinal distress (Rothan and Byrareddy, 2020). Data from faecal and respiratory specimen analyses suggest that SARS-CoV-2 virus particles survive longer in the gastrointestinal tract than in the respiratory tract (Tian et al., 2020). Moreover, SARS-CoV-2 RNA has been detected in feces of both symptomatic and asymptomatic infected individuals (Ahmed et al., 2020a). As such, the monitoring of wastewater for SARS-CoV-2 viral fragment occurrence to investigate the prevalence of COVID-19 infections in a given population has been gaining momentum as the pandemic continues to spread (Carducci et al., 2020; Foladori et al., 2020; Gonzalez et al., 2020; Kumar et al., 2020).

Reliable tests for SARS-CoV-2 infections target the viral genome through quantitative reverse transcription PCR (RT-qPCR) (Bustin and Nolan, 2020; Randazzo et al., 2020). The use of RT-qPCR led to the first report of SARS-CoV-2 RNA detection in untreated wastewater in a proof-of-concept study that demonstrated the applicability of wastewater surveillance for COVID-19 as a tool to understand infection rates within communities (Ahmed et al., 2020a). To effectively monitor SARS-CoV-2 viral fragment occurrence in wastewater, the development of an efficient and reliable methodology for viral fragment recovery from this complex matrix is paramount.

For most techniques, SARS-CoV-2 analysis in wastewater requires pre-concentration of the sample. Among the most common methods of virus concentration in wastewater are ultrafiltration, polyethylene glycol (PEG) precipitation, and ultracentrifugation (Ahmed et al., 2020c, 2020a; La Rosa et al., 2020; Medema et al., 2020; Randazzo et al., 2020; Wu et al., 2020; Wurtzer et al., 2020). Most of these RNA concentration methods are cost-ineffective, time-consuming, and labour-intensive (Ahmed et al., 2020c). Moreover, information on the recovery efficiencies of current methods for measuring SARS-CoV-2 in wastewater is limited (Ahmed et al., 2020c; Michael-Kordatou et al., 2020), as most studies use surrogates (e.g. murine hepatitis virus (MHV), MS2 (an F-specific RNA phage), and other coronaviruses), to perform recovery efficiency studies (Ye et al., 2016). Green et al., 2020 reported a 12% recovery of inactive SARS-CoV-2 from 20-mL samples of wastewater using ultracentrifugation.

In the last decade, several studies have demonstrated the use of magnetic beads to extract nucleic acids from a variety of matrices, including serum (Sun et al., 2014), urine (Shan et al., 2012), sputum (He et al., 2017), whole blood (Albertoni et al., 2011), tissue (Mathot et al., 2011), and wastewater (La Rosa et al., 2010; Yuan et al., 2019). While conventional approaches involve both pre-concentration and extraction using magnetic beads, an emerging methodology involves direct extraction using magnetic beads in the absence of a pre-concentration step. The advantages of direct RNA extraction via magnetic beads are that the procedure is rapid, cost-effective, and does not require the use of specialized equipment. Direct RNA extraction also increases viral recovery efficiency, as pre-concentration methods often significantly reduce overall recovery (Gonzalez *et al*., 2020), and allows for larger reaction volumes, potentially bridging the gap between pre-concentration sample volumes and traditional extraction sample volumes. Such advantages allow for greater diversity in monitoring applications, which is valuable for laboratories with limited resources. This study advances the application of magnetic bead extraction approaches through the development of a new method optimized for direct, rapid extraction of SARS-CoV-2 from wastewater. Here we report the recovery efficiency of the method using deionized water and municipal wastewater spiked with different viral surrogates and heat- and gamma-inactivated SARS-CoV-2.

## 2. MATERIALS AND METHODS

### 2.1 Reagents

Accuplex SARS-CoV-2 Positive Reference Material (ASCV-2) was purchased from Seracare Life Sciences Inc (Milford, MA, USA). Wastewater samples spiked with Gamma Inactivated SARS-COV-2/Canada/ON/VIDO-01/2020 and Human Coronavirus 229E were sourced from the National Microbiology Laboratory (Winnipeg, MB, Canada), Drs. Mubareka and Kozak at Sunnybrook Health Sciences Centre, University of Toronto, and VIDO-Intervac at the University of Saskatchewan. Deionized (DI) water was produced by a Milli-Q system (Reference A+, Millipore) and had a total organic carbon (TOC) concentration < 5 µg L^-1^ and a resistivity of 18.2 mΩ cm^-1^. Ethanol (EtOH) was purchased from Fisher Scientific (Ottawa, ON, CA). DL-Dithiothreitol (DTT, ∼1 M) was obtained from Sigma Aldrich (Ottawa, ON, CA). Mixed cellulose ester membranes (0.45-µm pore size, 47-mm diameter) were purchased from Merck Millipore Ltd. (Ottawa, ON, CA). Magnetic binding beads (20 g L^-1^), RNA isolation kits, and SARS-CoV-2 assay kits were obtained from LuminUltra Technologies Ltd (Fredericton, NB, CA).

### 2.2 Wastewater sample collection and preparation

Wastewater samples were collected from three communities in Atlantic Canada: Community A (grab sample from one sewershed site and influent 24-h composite samples from two WWTFs), Community B (grab samples from the pump station and influent and effluent of the WWTF [two-cell aerated lagoon with disinfection using chlorine gas]), and Community C (sewershed grab samples at two locations and WWTF influent 24-h composite samples). Samples were collected in volumes of at least 100 mL and transported to Dalhousie University on ice. For each sample, an aliquot of 45-mL was transferred to a 50-mL centrifuge tube and kept at 4 °C for up to 24 h. The wastewater aliquot was seeded with Accuplex SARS-CoV-2 Positive Reference Material to yield a concentration of 1×10^6^ gene copies L^-1^ (GU L^-1^). The spiked sample was mixed thoroughly and incubated at 4 °C for 30 min prior to RNA extraction. All samples were stored overnight at 4 °C and processed the next morning for RNA extraction and RT-qPCR analysis.

Additional wastewater samples were provided by the Winnipeg Wastewater Treatment Plant (WWTP) (Winnipeg, Manitoba, Canada) to assess the RNA extraction method for the recovery and quantification of SARS-CoV-2 surrogates spiked at different concentrations. The grab wastewater samples were spiked with high (1.8×10^6^ ± 2.0 x10^5^ GU L^-1^) and low (1.8×10^4^ ± 2.0 ×10^3^ GU L^-1^) concentrations of gamma-irradiated SARS-CoV-2 NR-52287 (GI-SCV-2) and high (1.0×10^6^ infectious units L^-1^) and low (1.0×10^4^ infectious units L^-1^) concentrations of Human Coronavirus 229E ATCC^®^ VR-740^™^ (HCV 229E). Viral surrogates were obtained from the University of Alberta and were spiked at the National Microbiology Laboratory. The seeded wastewater was shipped to Dalhousie University on ice, and the samples were analysed within 48 h of seeding. The wastewater was kept at 4 °C during transport and throughout the preparation of spiked samples.

### 2.3 Determination of RNA extraction conditions with magnetic beads using a factorial design

A screening statistical design was chosen to establish optimal conditions for the RNA extraction of ASCV-2 from wastewater and maximum recovery efficiency for the RT-qPCR. Thus, a full-factorial design of four factors was used to assess the magnetic bead-based extraction protocol in RNA extraction. All experiments were performed according to the experimental design shown in Table 1. A total of 16 experimental runs were carried out randomly to limit the occurrence of potential bias. The four independent variables in this study were ASCV-2 spiked sample volume (mL) (*x*_*1*_), concentration of magnetic beads (µL mL^-1^ sample) (*x*_*2*_), RNA elution temperature (°C) (*x*_*3*_), and water matrix (DI water and wastewater) (*x*_*4*_). The dependent variable was evaluated as ASCV-2 recovery efficiency (%), calculated based on the number of copies quantified for each experimental run by RT-qPCR using Equation 1:

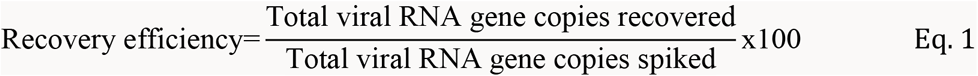

**Table 1.** Experimental factors and their levels for the 2^4^ full-factorial design.

| Factor | Level |  |
| --- | --- | --- |
|  | -1 | +1 |
| $x_1$: Spiked water volume | 1 mL | 2.5 mL |
| $x_2$: Magnetic bead concentration | 15 $\mu\text{L}$ per mL sample | 100 $\mu\text{L}$ per mL sample |
| $x_3$: Elution temperature | 30 $^{\circ}\text{C}$ | 60 $^{\circ}\text{C}$ |
| $x_4$: Water matrix | DI water | Wastewater |

Each seeded sample was prepared by spiking ASCV-2 in either DI water or wastewater to a concentration of 1×10^6^ GU L^-1^, as described in section *2*.*2 Wastewater sample collection and preparation*. All RNA extractions were performed with reagents provided by LuminUltra Technologies Ltd (Fredericton, NB, CA). In a 15-mL centrifuge tube, 4.9 or 6.5 mL of Lysis buffer was added to 1 or 2.5 mL of spiked water sample, vortexed for 30 sec, and immediately incubated at 30 °C for 10 min. A volume of 3.5 mL EtOH was added to the lysed sample; the tube was gently inverted several times to mix thoroughly and then spiked with binding beads at a concentration of either 15 or 100 µL per mL of sample. The mixture was vortexed for 30 sec and incubated again at 30 °C for 10 min. The magnetic beads were precipitated by applying a magnet, and the supernatant was discarded. The magnetic beads were transferred to a 2-mL microcentrifuge tube and washed three times with 1 mL of Wash I solution. A second wash step using 1 mL of Wash II solution was carried out three times. For each wash, the magnetic beads were vortexed for 30 sec, and the supernatant was discarded after magnet precipitation of the beads. Once washed, the tubes containing the magnetic beads were placed at room temperature with the caps off for approximately 1 h to evaporate residual EtOH. Then, 50 µL of elution buffer (preheated to 30 or 60 °C) was added to the magnetic beads. The tube was vortexed for 30 sec and incubated at either 30 or 60 °C for 5 min. Finally, the magnet was applied to ensure separation, and the elution buffer was collected into a sterile tube for analysis. Controls without seeded ASCV-2 were prepared to identify background concentrations of the virus in wastewater. A graphical representation of the RNA extraction protocol using magnetic beads is shown in Figure 1.

**Figure 1.**
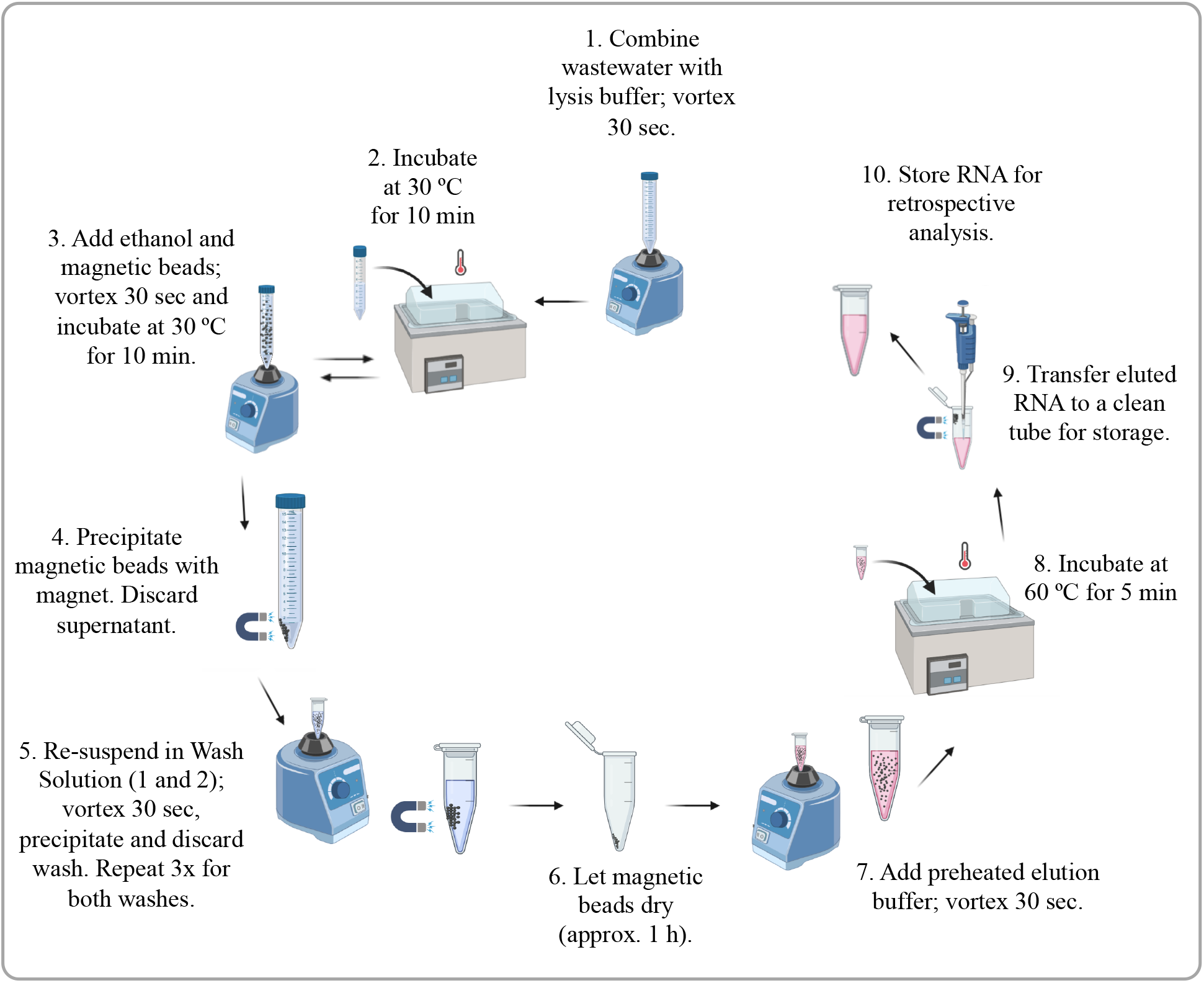
Conceptual framework for the extraction of RNA from wastewater using magnetic beads. Created with BioRender.com.

### 2.4 Evaluation of RNA extraction method for recovery using gamma-irradiated SARS-CoV-2 and Human Coronavirus surrogates in wastewater

Optimal conditions for RNA extraction using magnetic beads identified with the factorial design were evaluated for the recovery and quantification of SARS-CoV-2 surrogates. A grab sample from the Winnipeg WWTP was obtained, and aliquots were individually spiked with high and low concentrations of both GI-SCV-2 and HCV 229E at the National Microbiology Laboratory (as described in section 2.2 *Wastewater sample collection and preparation*). A total of three aliquots of 100 mL for each surrogate spike condition were prepared. Moreover, a negative control was added to assess the background concentration of both surrogates in the wastewater. For RNA extraction, each of the aliquots for every surrogate spike condition was processed in duplicate with the optimal parameters of 1 mL sample volume, 100 µL of magnetic beads per mL of sample, and RNA elution temperature of 60 °C. Following extraction, RNA was quantified by RT-qPCR and the recovery efficiency of each surrogate spike condition was calculated.

### 2.5 Assessing matrix effects of wastewater on RNA recovery

The effect of the wastewater matrix in the magnetic bead-based extraction protocol was studied to improve RNA recovery efficiency. Wastewater seeded with ASCV-2 was processed using two different strategies: filtration pre-treatment and addition of DTT (∼1 M) to the lysis buffer. For the first approach, wastewater was pre-treated by filtering through a 0.45-µm pore membrane to remove particulates that may saturate the magnetic beads. A sample spiked with 1×10^6^ GU L^-1^ of ASCV-2 was passed through a 47-mm diameter mixed cellulose ester membrane via a sterile magnetic filter funnel (Pall Corporation, Mississauga, Ontario, Canada) and filtration flask. The filtrate was immediately collected into a sterile conical tube and used for RNA extraction. The second strategy involved the addition of DTT (∼1 M) during the lysis step to inactivate RNases that may degrade RNA. DTT was added to the lysis buffer and spiked wastewater (filtered and non-filtered) at a concentration of 20 mM.

A total of four experiments were performed in triplicate to assess the effect of wastewater pre-filtration and addition of DTT on RNA recovery efficiency using the magnetic bead-based extraction protocol. Viral RNA extraction and quantification was carried out using the optimal parameters identified by the experimental design: 1 mL sample volume, 100 µL of magnetic beads per mL of sample, and RNA elution temperature of 60 °C.

### 2.6 RT-qPCR assay

RNA samples were processed by RT-qPCR on a GeneCount^®^ Q8 instrument (LuminUltra Technologies Ltd, Fredericton, CA). The sequences for primers and probes published by US CDC used in this study are shown in Table 2 (CDC, 2020). For the detection of SARS-CoV-2, the RT-qPCR amplifications were performed in 20-µL reactions using the GeneCount SARS-CoV-2 Screening kit (LuminUltra Technologies Ltd, Fredericton, CA), which contained 15 µL of Master Mix and 5 µL of template RNA. The SARS-CoV-2 nucleocapsid gene DNA plasmid was used as a positive control. Thermal cycling reactions were carried out as follows: a pre-denaturation step at 55 °C for 10 min followed by a second pre-denaturation step at 95 °C for 1 min; 45 cycles of 95 °C for 10 sec and 55 °C for 45 sec; and a final hold step at 50 °C for 1 min. Reactions were considered positive when cycle threshold (Ct) values were below 40. The upper Ct value detection threshold for the RT-qPCR was 40 cycles corresponding to 1.4 copies per reaction.

**Table 2.**
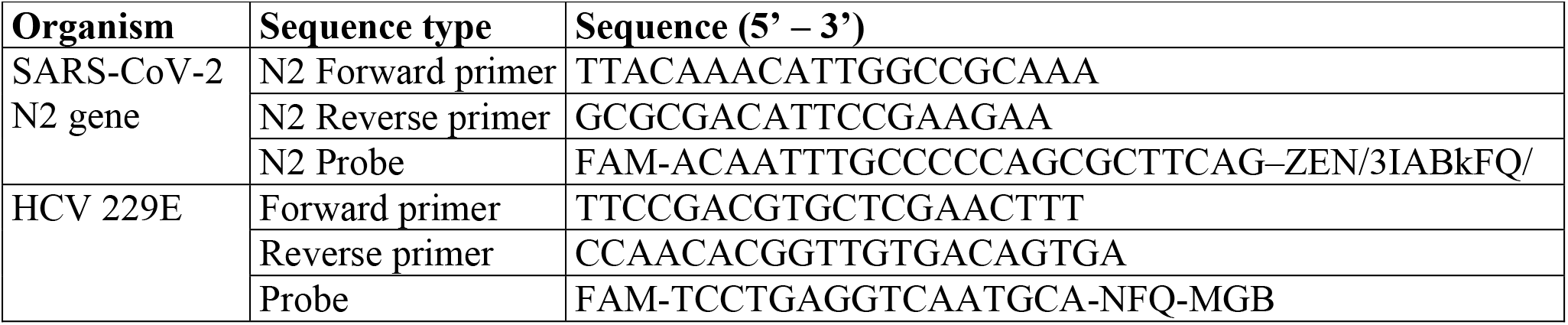
Sequences for primers and probes of viral surrogates used in this study.

For HCV 229E, a previously published Taq-Man based RT-qPCR assay was used (Vijgen et al., 2005). HCV 229E RT-qPCR analyses were performed in 10-µL reaction mixtures using TaqMan^™^ Fast Virus one-step Master Mix (Thermo Fisher Scientific, CA). The reaction mixture contained 2.5 µL of Master Mix, 10 µM of forward primer, 10 µM of reverse primer, 10 µM of probe, 1.5 µL of PCR grade water, and 5 µL of template RNA. Thermal cycling conditions consisted of RT at 50 °C for 5 min, enzyme activation at 95 °C for 20 sec, and 45 cycles of 95 °C for 3 sec followed by 60 °C for 30 sec. All RT-qPCR analyses were carried out at least in duplicate, and for each qPCR run, both a positive and negative control were included.

### 2.7 Method Limit of Detection (MLOD) determination

The method limit of detection (MLOD) of the optimized RNA extraction method was calculated using ASCV-2 spiked into wastewater that was pre-filtered as described in Section 2.5 *Assessing matrix effects of wastewater on RNA recovery*. The synthetic RNA was seeded into pre-filtered wastewater in a series of decreasing concentrations (1×10^3^ to 1×10^6^ GU L^-1^). All diluted solutions were subjected to the optimized RNA extraction method and assessed by RT-qPCR in six replicates. Before adding ASCV-2, the wastewater sample was analyzed for the presence of SARS-CoV-2 to obtain background levels. The percentage of replicates that resulted in a positive detection for each dilution were compared to the spiked virus concentration, and the data were fitted to a Gompertz model (Tjørve and Tjørve, 2017). Using this model, the dilution reporting the lowest non-negative concentration at a 95% confidence level was selected as the MLOD.

### 2.8 Application of the RNA extraction method for the detection and quantification of SARS-CoV-2 in wastewater

The application of the RNA extraction and RT-qPCR methods were evaluated in raw wastewater samples from three locations in Atlantic Canada. In Fall 2020, wastewater samples were collected from Communities A, B, and C over five, eight, and four sampling events, respectively. RNA fragment extraction was carried out using the optimal parameters identified by the experimental design: 1 mL sample volume, 100 µL of magnetic beads per mL of sample, and RNA elution temperature of 60 °C.

### 2.9 Quality control

To minimize contamination, RNA extraction and RT-qPCR assays were carried out in separate laboratories. A method blank was included during RNA extraction to account for any contamination during sample processing. The extracted RNA was stored at –76 °C and subjected to RT-qPCR within the same day of extraction.

### 2.10 Statistical analysis

All analyses were performed at least in duplicate. Results were expressed as average values ± standard deviation and were submitted to analysis of variance (ANOVA) with a 95% confidence level. The response variable in the full-factorial design was modeled as a linear combination of the main effects and two-way interactions, and residuals were approximately Gaussian and homoscedastic. For experiments assessing the effect of wastewater matrix on recovery efficiency, Tukey’s test for post-hoc analysis was used to evaluate group pairings that were significantly different. The MLOD was assessed by plotting percent positive results against the known spiked ASCV-2 concentrations in the series of MLOD test wastewater samples. A best fit Gompertz model was fitted to the data, and the MLOD was calculated from the equation generated from the best fit model when the value of percent positive (y-axis) was 95%. All statistical analyses were performed using R studio software v. 3.6.2 (‘easynls’, ‘ggplot2’, ‘tidyverse’ packages) (Arnhold, 2017; R Core Team, 2020; Wickham et al., 2019).

## 3 RESULTS AND DISCUSSION

### 3.1 Effects of RNA extraction parameters on recovery efficiency

The effects of sample volume, concentration of magnetic beads, and elution temperature on ASCV-2 RNA recovery in DI water and municipal wastewater were assessed through a factorial experimental design (Table 3). The combination of optimized method parameters from the full-factorial design that resulted in the highest RNA recovery in both DI water (26.0 ± 0.8%) and wastewater (11.8 ± 1.4%) was a sample volume of 1.0 mL, a magnetic beads concentration of 100 µL mL^-1^ sample, and an elution temperature of 60 °C. Overall, ASCV-2 recoveries were clearly influenced by the water matrix with values ranging from 1.3 to 11.8% for wastewater and 6.3 to 26.0% for DI water. To better visualize the influence of the factors on RNA recovery, a response surface contour plot illustrating the relationship among the dependent (sample volume, bead mix concentration, elution temperature, and water matrix) and independent (% recovery) factors was generated (Figure 2). It is important to note that SARS-CoV-2 was not detected in any of the unseeded wastewater samples.

**Table 3.**
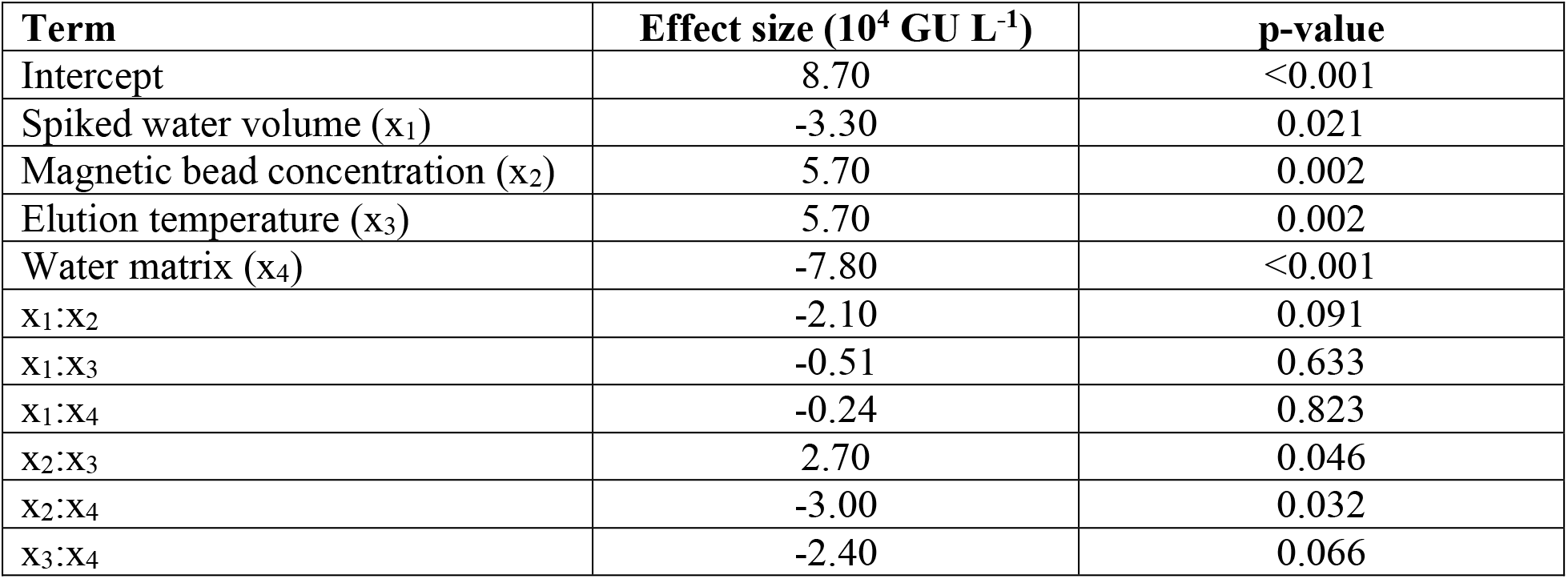
A summary of the linear model, which explained 97% of the variation in recovery. Effect estimates (other than the intercept) are twice their corresponding linear regression coefficients.

| Term | Effect size ($10^4$ GU L<sup>-1</sup>) | p-value |
| --- | --- | --- |
| Intercept | 8.70 | <0.001 |
| Spiked water volume ($x_1$) | -3.30 | 0.021 |
| Magnetic bead concentration ($x_2$) | 5.70 | 0.002 |
| Elution temperature ($x_3$) | 5.70 | 0.002 |
| Water matrix ($x_4$) | -7.80 | <0.001 |
| $x_1:x_2$ | -2.10 | 0.091 |
| $x_1:x_3$ | -0.51 | 0.633 |
| $x_1:x_4$ | -0.24 | 0.823 |
| $x_2:x_3$ | 2.70 | 0.046 |
| $x_2:x_4$ | -3.00 | 0.032 |
| $x_3:x_4$ | -2.40 | 0.066 |

**Figure 2.**
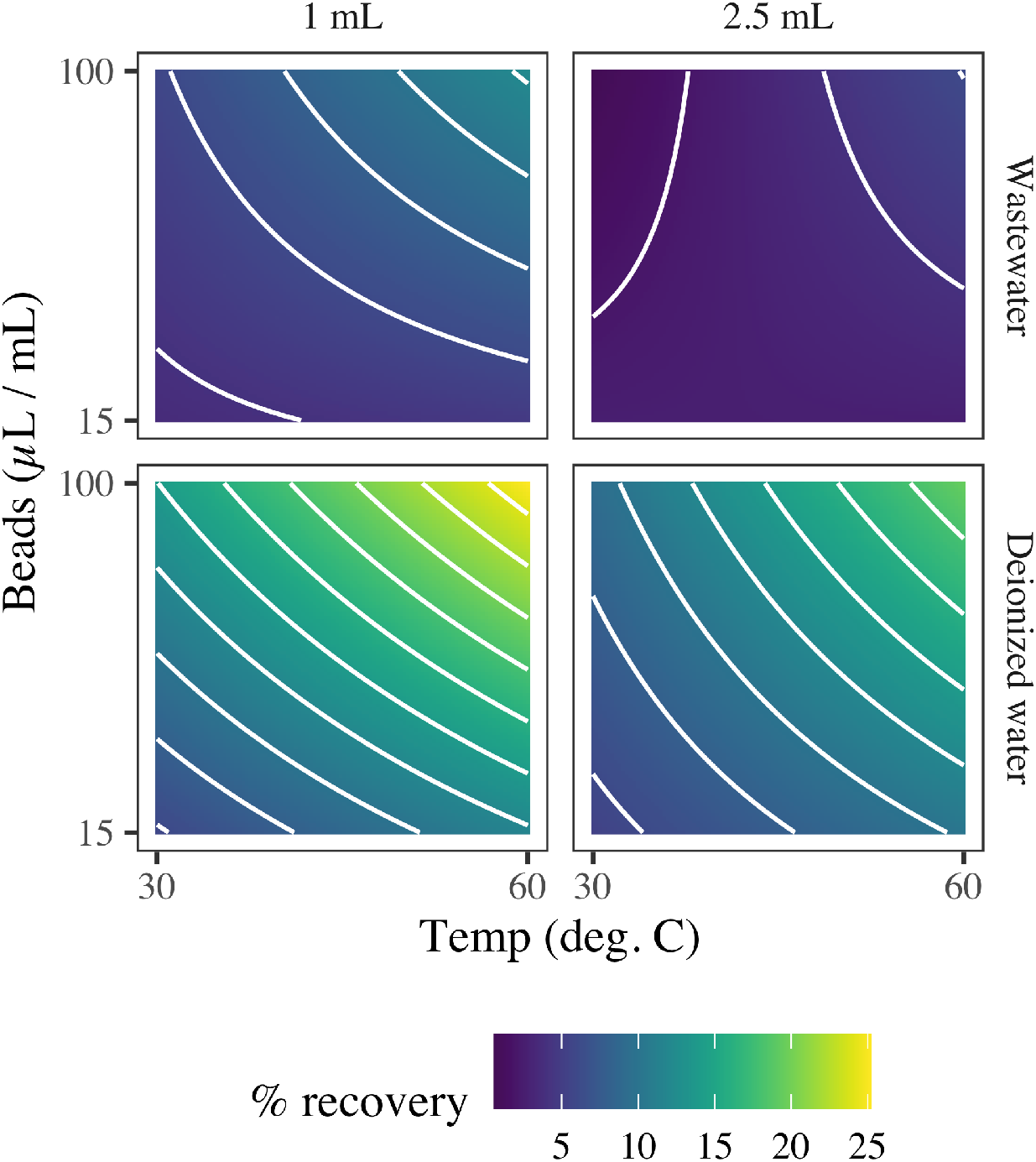
Response surface contour plot illustrating the effect of sample volume, bead mix concentration, elution temperature, and water matrix (and their two-way interactions) on percent recovery.

The factorial experimental design demonstrated that a lower sample volume of 1 mL resulted in consistently higher mean RNA recoveries than did a sample volume of 2.5 mL when assessed at the same conditions for all other variables. The effect calculated from the linear model showed that increasing the volume from 1 to 2.5 mL decreased recovery by 3.3×10^4^ GU L^-1^. These results suggest that a larger volume may have diluted the concentration of magnetic beads and decreased contact between the beads and RNA. Thus, using smaller sample volumes allowed more bead-RNA collisions and subsequent binding of RNA onto the magnetic beads, which was reflected in higher recovery yields (Yuan et al., 2019).

In recent comparable wastewater studies, SARS-CoV-2 RNA was concentrated from considerably larger sample volumes. For instance, SARS-CoV-2 RNA was detected in all of 54 untreated Detroit wastewater samples following RNA concentration from 45-L sample volumes (Miyani *et al*., 2020). Ahmed *et al*., (2020b) reported “small volumes” of wastewater for RNA extraction as samples consisting of 50 mL, which is 50-fold greater than the optimal volume determined in this study. Working with smaller wastewater sample volumes is advantageous, as they are easier and safer to handle, store, and transport. While this method performs optimally at low sample volumes (1 mL), it does not incorporate a concentration step, indicating that it is suitable for wastewater samples that contain SARS-CoV-2 in concentrations present above its method detection limit.

Pre-concentration of RNA in wastewater does not necessarily result in a higher recovery efficiency. Gonzalez *et al*., (2020) compared the recovery efficiency of Bovine coronavirus (BCoV) and bovine respiratory syncytial virus (BRSV) from concentrated and unconcentrated wastewater at a sample volume of 2 mL. Two concentration techniques were used: concentrating pipette with centrifugation and electronegative filtration. Interestingly, recoveries of the surrogates without concentration were 59 and 75% for BCoV and BRSV, respectively, while neither concentration technique resulted in recoveries above 7.6% for either surrogate. This reduced recovery observed by Gonzalez *et al*., (2020) was attributed to numerous workflow concentration steps.

The concentration of magnetic beads added to the sample also greatly impacted RNA recovery, as the beads play a primary role in the binding of target nucleic acids. Certainly, a greater concentration of magnetic beads provides additional surface area, allowing for more binding sites for nucleic acids (Yuan et al., 2019). A magnetic beads concentration of 100 µL mL^-1^ sample resulted in significantly higher RNA recovery efficiencies (about 60 and 70% increases in recovery from wastewater and DI water, respectively) than did a concentration of 15 µL mL^-1^ sample. Furthermore, when magnetic beads were dosed at a concentration of 100 µL mL^-1^ sample, the recovery efficiency was maximized at low sample volume and minimized at higher sample volumes. The effect of this factor calculated from the linear model showed that increasing the concentration of magnetic beads from 15 to 100 µL mL^-1^ sample increased recovery by 5.7×10^4^ GU L^-1^. These results support our initial premise that a larger sample volume may dilute the concentration of magnetic beads, which leads to lower RNA recoveries. In agreement with our observations, Yuan et al., (2019) found that although smaller sample volumes contain less total nucleic material, increased collisions between magnetic beads and target nucleic acids produced higher recoveries, while larger sample volumes diluted the concentration of magnetic beads and impaired eDNA recovery.

In both matrices, increasing the elution temperature from 30 to 60 °C resulted in a 60% greater RNA recovery. The effect of elution temperature calculated from the linear model showed that an increase from 30 to 60 °C increased recovery by 5.7×10^4^ GU L^-1^. Previous studies have outlined the important role of temperature for the desorption of nucleic acids (Elaissari et al., 1999; Li et al., 2011, 2012). Nonetheless, there is a lack of consensus as many methods are performed at room temperature (He et al., 2017; La Rosa et al., 2010; Sun et al., 2014), whereas other studies have demonstrated that more RNA is released at increased temperatures. For example, high temperatures have been reported to weaken the hydrogen bonds formed between nucleic acids-adsorption materials, therefore, serving as an ideal tool to enhance the elution process (Li et al., 2011). However, it should be considered that high elution temperatures (>60 °C) may increase the risk of RNA denaturation. In our preliminary experiments, recovery efficiency increased considerably when RNA was eluted at 55 °C in comparison to room temperature (data not shown), and it was further improved by incubating at 60 °C. It is likely that the short incubation periods used in this work in combination with the pre-heated elution buffer promoted the solubilization of extracted RNA while minimizing RNA degradation.

Of all four factors assessed in the factorial design recovery experiments, the matrix showed the largest impact on RNA recovery. The effect of the matrix calculated from the ANOVA showed that extracting RNA from wastewater (rather than DI water) decreased recovery by 7.8×10^4^ GU L^-1^. The higher recovery yields obtained with DI water demonstrated the effect of the complex composition of wastewater on the performance of the magnetic bead-based extraction protocol. RNA recovery efficiency can be influenced by the matrix when the particulate and dissolved components in the wastewater are carried along with the target virus during isolation (Michael-Kordatou et al., 2020). As RNA extraction was performed for raw wastewater without any pre-treatment, it is likely that debris and solids present in the matrix competed with nucleic acids for binding sites on the magnetic beads (Huggett et al., 2008). Due to this reduced surface area, lower quantities of RNA were able to bind to the magnetic beads in the wastewater than in DI water.

In addition to the influence of physical interactions, the impact of the matrix on the performance of the magnetic beads and virus recovery may also have a biochemical nature. It is well known that the large variety of organic and inorganic molecules in wastewater can inhibit the RT-qPCR process by reducing its sensitivity. These compounds, commonly referred to as PCR inhibitors (Acharya et al., 2017), may be co-concentrated with the virus during the processing of large sample volumes for the detection of low viral concentrations (Ahmed et al., 2020d). In this work, the degree of inhibition was expected to be lower, as samples were not pre-concentrated prior to RNA extraction. Nonetheless, inhibition was assessed by comparing Ct values obtained for wastewater samples to those obtained for DI water samples (reference value). Samples having Ct values greater than two cycles above the reference point were considered to be impacted by PCR inhibition (Ahmed et al., 2020b). In these experiments, the difference between the wastewater Ct value and the reference value for DI water was less than two cycles, which suggests low inhibition from the wastewater matrix. These results may be attributed to small sample volumes used for RNA extraction (1 to 2.5 mL). For example, Hata et al. (2011) observed that the use of larger sample volumes (8.0 to 200 L) was associated with a greater inhibitory effect. Conversely, other studies have observed no trends in PCR inhibition with respect to sample volume for SARS-CoV-2 using RT-qPCR (Gibson et al., 2012; Medema et al., 2020). As such, further research is necessary to elucidate the effect of wastewater matrix on the performance of magnetic beads to enhance the recovery of ASCV-2.

### 3.2 Minimizing effects of the wastewater matrix on RNA recovery

The results for assessing the impacts of pre-filtration and the addition of DTT on wastewater matrix effects are shown in Table 4. Treatment A (unfiltered wastewater without DTT) provided the highest mean ASCV-2 recovery of 17.4%. The second highest mean recovery (16%) was for treatment C, filtered wastewater without DTT. Overall, the addition of DTT in treatments B and D showed similar results with slightly lower mean recovery yields compared to those obtained with unfiltered and filtered wastewater only. Tukey’s test for post-hoc analysis showed that the mean recovery efficiency of treatment A was significantly different than that of treatment D (*p* < 0.05).

**Table 4.** Effects of wastewater pre-filtration and DTT addition on recovery efficiency of ASCV-2 using the magnetic bead-based extraction protocol (n=3).

| Sample treatment | Mean ± SD of % recovery of ASCV-2 |
| --- | --- |
| A. Wastewater | 17.4 ± 0.4 |
| B. Wastewater + DTT | 14.0 ± 1.7 |
| C. Filtered wastewater | 16.0 ± 0.9 |
| D. Filtered wastewater + DTT | 14.2 ± 2.4 |

The complex diversity in chemical and biological composition of wastewater can impede accurate detection of viruses in these samples. In addition to high suspended solids and organic matter concentrations, raw wastewater usually contains compounds such as polysaccharides, metal ions and RNases which are common PCR inhibitors (Corpuz et al., 2020; Michael-Kordatou et al., 2020; Schrader et al., 2012). Particularly, the abundance of RNases can significantly compromise the stability of RNA throughout the extraction and purification steps. As RNase activity requires its disulfide bond to be intact, denaturation of the enzyme can be achieved through addition of reducing agents, such as DTT, during lysis (Lu and Chang, 2010). In this study, the recovery of ASCV-2 was not improved with the addition of DTT to either filtered or unfiltered wastewater. It is possible that the concentration of DTT (20 mM) spiked into the lysis buffer was insufficient to reduce the disulfide bonds in RNase, as previous investigations have shown that concentrations of 40 to 100 mM led to the effective inactivation of these proteins (Lu and Chang, 2010; Rogacs et al., 2012). However, in a recent study that evaluated the reproducibility and sensitivity of 36 methods to quantify SARS-CoV-2 in raw wastewater, neither the removal of solids from the sample nor the addition of a concentration step demonstrated a clear impact on recovery efficiency (Pecson et al., 2020).

### 3.3 Recovery of gamma-irradiated SARS-CoV-2 and HCV 229E surrogates in wastewater

The optimized magnetic bead-based extraction protocol was evaluated for recovery of GI-SCV-2 and HCV 229E as SARS-CoV-2 surrogates in wastewater. Overall, the method allowed differentiation between high and low concentrations, and non-spiked wastewater samples. Neither of the two surrogates were detected in the negative samples. Mean concentrations of 8.33×10^4^ ± 7.8×10^3^ GU L^-1^ and 1.55×10^4^ ± 3.6×10^3^ GU L^-1^ were recovered for samples spiked at high and low concentrations of GI-SCV-2.

In preliminary experiments, mean GI-SCV-2 recovery was 86.1% for the low spike concentration, whereas at the high spike concentration, mean recovery dropped to 4.6%. A potential cause for the decrease in GI-SCV-2 recovery at the high spike concentration may be PCR inhibition. Although the use of magnetic beads has been expected to counter inhibitory effects better than other column-based extraction methods (Hata et al., 2011), they are still affected by inhibitory compounds. A common approach to reducing inhibitor interference is through RNA sample dilutions. However, this results in direct dilution of both the inhibitory substances present in the sample (Michael-Kordatou et al., 2020) and target RNA. Moreover, dilution of samples containing low levels of RNA may cause target RNA concentrations to fall below the LOD. Although dilution reduces the overall RNA concentration in the sample, the simultaneous dilution of inhibitors may increase the efficacy of the PCR reaction (Wang et al., 2017), thus resulting in a higher concentration of RNA detected.

To investigate inhibition as a potential cause of the GI-SCV-2 recovery loss, RNA extracts from the wastewater samples spiked with 1.8×10^4^ and 1.8×10^6^ GU L^-1^ were diluted 10- and 50-fold, respectively (n=3). For the low spike concentration, the RNA was diluted beyond the detection limit and could not be enumerated. However, recoveries for the diluted RNA extracts from the high spike concentration samples increased from 4.6 to approximately 100.1%. This observation supports the hypothesis that the wastewater samples contained PCR inhibitors that were co-extracted with surrogate RNA. During qPCR analysis, these inhibitors likely interfered with the accurate amplification of RNA samples and substantially reduced the recovery of GI-SCV-2. When the extracted RNA was diluted (along with the co-extracted inhibitors) and reanalyzed, the diminished interference in the qPCR reaction allowed for more accurate RNA quantitation.

For the recovery of HCV 229E, Ct values from the qPCR were used to provide an indication of the copy number of target RNA. Logarithmically, Ct values are inversely proportional to viral load. Consequently, as cycle numbers increase so do undesirable PCR products. Thus, Ct values are bounded by a range of positive and negative cut-off values (Public Health Ontario and The Ontario COVID-19 Testing Technical Working Group, 2020). The positive cut-off, determined by manufacturers or laboratories during method validation, is designated as the Ct value that represents the lowest copies of target RNA that can be reliably detected (e.g., Ct ≤ 38). A negative cut-off represents a Ct value at which the target RNA can no longer be detected. Ct values for HCV 229E ranged from 32.90 to 35.21 for the wastewater spiked with 1.0×10^6^ infectious units L^-1^ and from 37.42 to 38.22 for samples seeded with 1.0×10^4^ infectious units L^-1^.

### 3.4 Method limit of detection (MLOD)

The MLOD was evaluated by spiking ASCV-2 in filtered wastewater over a concentration range of 1×10^3^ to 1×10^6^ GU L^-1^ and calculated using a best fit Gompertz model (R^2^ = 0.980) (Figure 3). The method was able to detect 100% of the replicates when the virus was present at high concentrations (4.0×10^4^ to 1.0×10^6^ GU L^-1^). However, the percentage of positive replicates decreased as the ASCV-2 concentration decreased. As shown in Figure 3, the experimentally determined MLOD for the detection of ASCV-2 in wastewater with 95% confidence was approximately 4.6×10^4^ GU L^-1^. This value is within the range of theoretical limits of detection (10^3^ to 10^6^ GU L^-1^) reported for 36 different methods used to quantify SARS-CoV-2 in raw wastewater (Pecson et al., 2020).

**Figure 3.**
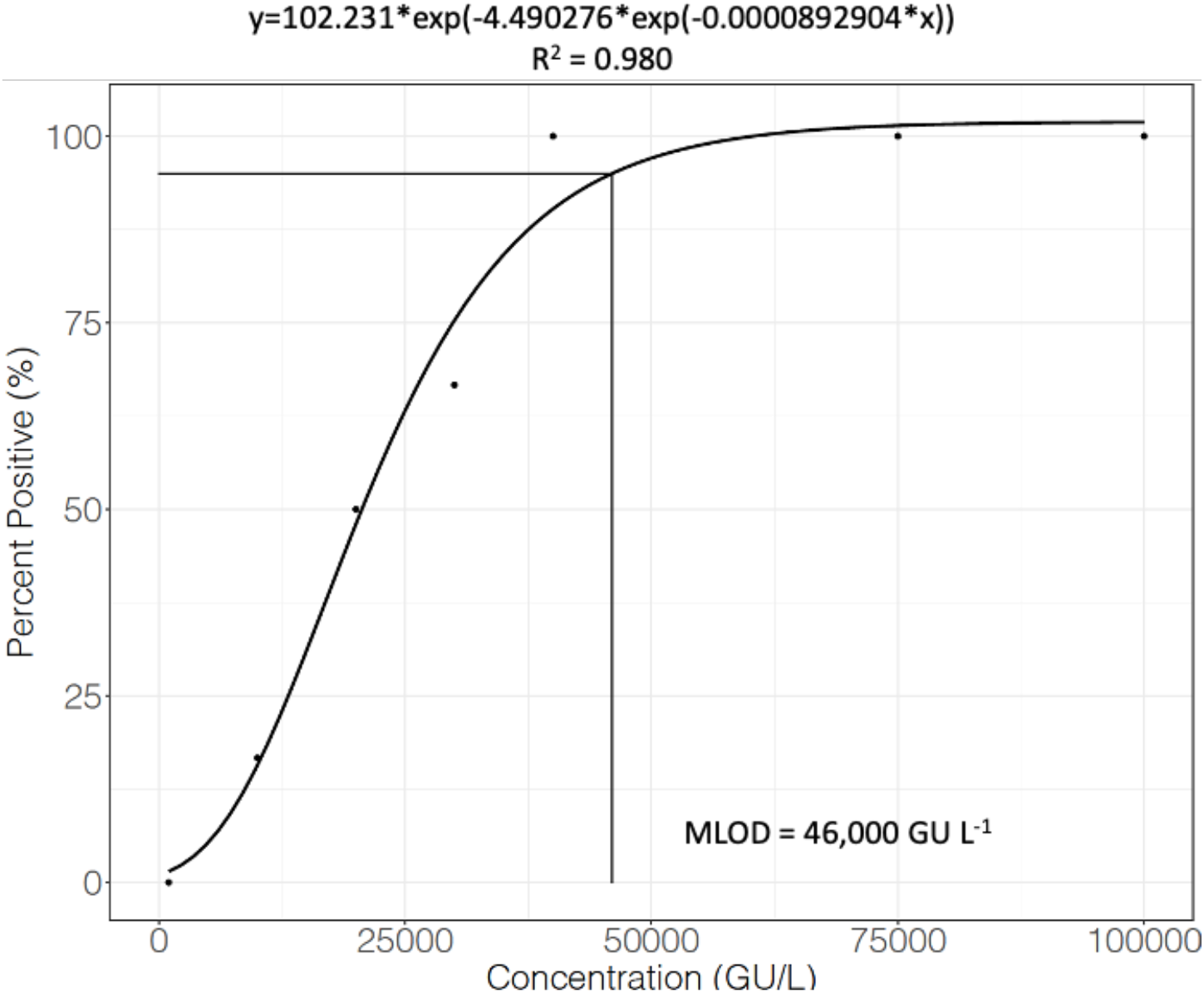
Calculation of the method limit of detection (MLOD) using a best fit Gompertz model (R^2^ = 0.980).

### 3.5 Detection of SARS-CoV-2 RNA in wastewater

The results of wastewater analysis for Communities A, B, and C are shown in Figure 4. A total of eight samples from the WWTFs (influent) in Community A were tested, none of which tested positive for SARS-CoV-2 RNA fragments. However, of the five samples analyzed from the sewershed in Community A, one tested positive for SARS-CoV-2 at a concentration of 3.25×10^2^ GU mL^-1^. Although this value is below the MLOD, the detection of SARS-CoV-2 in this sample may attributed to a difference in the degree of water matrix interferences. All 22 samples tested from the Community B WWTFs (influent/effluent) and pump station tested negative for SARS-CoV-2 RNA fragments. The presence of SARS-CoV-2 RNA was verified in samples collected from the Community C sewershed on Day 1 (6.6×10^4^ GU L^-1^) and Day 3 (1.4×10^4^ GU L^-1^) (Table 5).

**Table 5.** Detection of SARS-CoV-2 RNA in wastewater samples at a sewershed and WWTF in Community C.

| Sampling day | Mean SARS-CoV-2 RNA concentration obtained ($\text{GU L}^{-1}$) | |
| --- | --- | --- |
|  | Sewershed | WWTF Influent |
| Day 1 | $6.6 \times 10^4$ | ND |
| Day 3 | $1.4 \times 10^4$<sup>a</sup> | ND |
| Day 7 | ND | ND |
| Day 10 | ND | ND |

ND: Not detected
<sup>a</sup> Detected at Cycle threshold (Ct) = 40.

**Figure 4.**
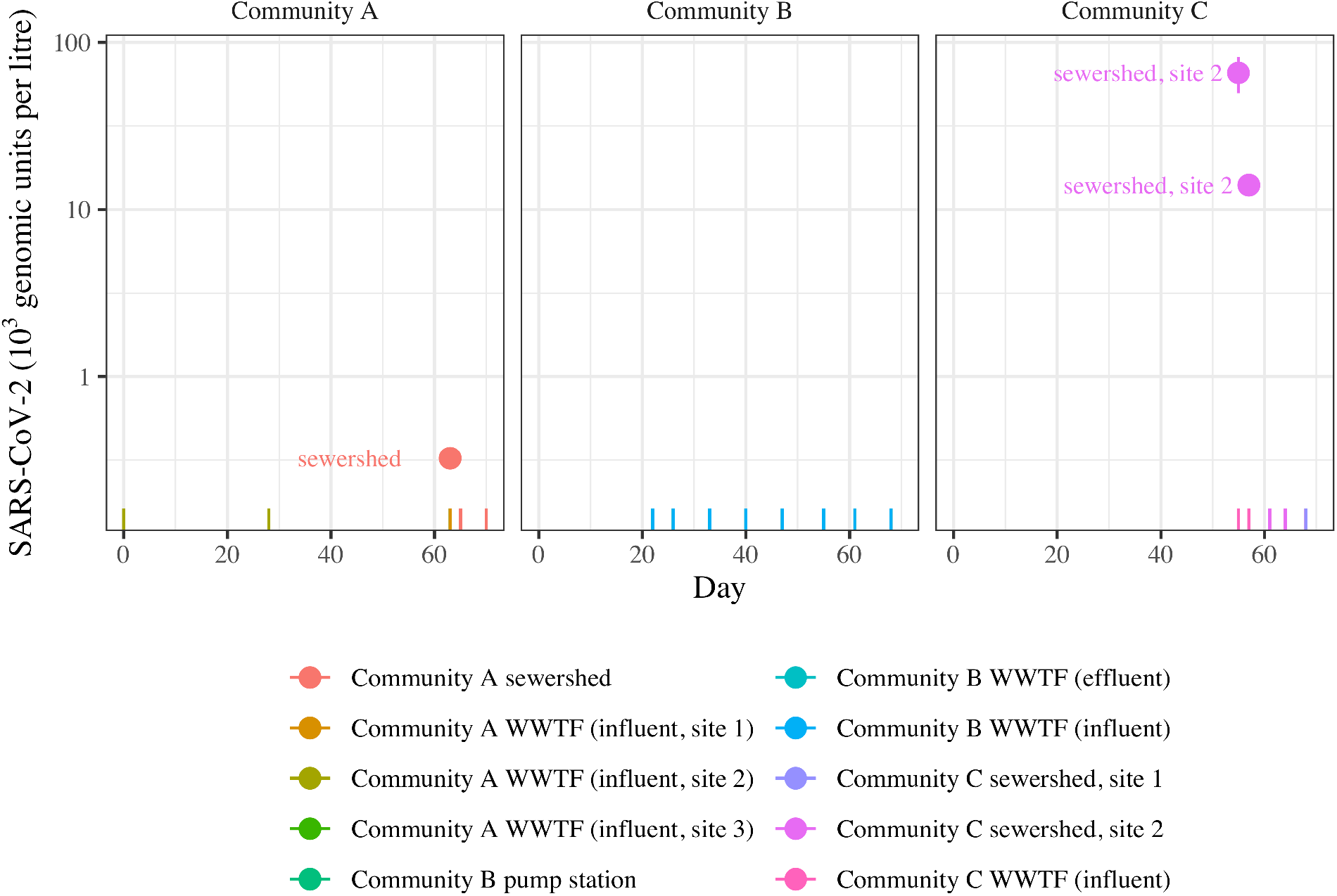
SARS-CoV-2 viral fragment (RNA) detections in wastewater in Communities A, B, and C (log scale). Error bars span one standard deviation about the mean; vertical lines at the bottom of each panel denote non-detects.

These results demonstrate that the magnetic bead-based extraction protocol allows detection of SARS-CoV-2 RNA fragments when the virus is present in the wastewater sample above the MLOD. However, the virus may remain undetected in samples processed using this method if the viral RNA is present in concentrations below the MLOD. As observed in Table 5, the concentration of SARS-CoV-2 remained below the MLOD during the third and fourth sampling events for the sewershed and for all sampling events for the WWTF Influent. Viral load from faeces varies largely within wastewater depending on the time of infection onset. Peak viral shedding for SARS-CoV-2 is known to occur within two to three weeks of the onset of initial symptoms (Zheng et al., 2020). The results presented in this section demonstrate the potential of the rapid method proposed in this work for the detection and surveillance of SARS-CoV-2 in wastewater using optimal method parameters: 1 mL sample volume, 100 µL of magnetic beads per mL of sample, and RNA elution temperature of 60 °C.

## 4. CONCLUSIONS

This work outlines the development and optimization of a rapid and effective magnetic bead-based extraction protocol in the detection of SARS-COV-2 in complex wastewater samples. Through the optimization of the developed method, we found that the water matrix impacted the recovery of ASCV-2 with values ranging from 1.3 to 11.8% for wastewater and 6.3 to 26.0% for DI water. Moreover, extracting RNA from wastewater (rather than DI water) decreased RNA recovery by 7.8×10^4^ GU L^-1^. Additional experiments testing the effects of filtration and the addition of DTT on RNA recovery showed that there was no significant difference in the recovery efficiency of ASCV-2 from filtered and non-filtered wastewater nor with the addition of DTT (20 mM) to the lysis buffer, and thus, further work may be necessary to address water matrix effects.

Independent of the water matrix, a higher elution temperature resulted in higher RNA recoveries in both DI water and wastewater, and recovery increased by 5.7×10^4^ GU L^-1^ as elution temperature increased from 30 to 60 °C. Moreover, a smaller sample volume of 1 mL resulted in higher mean RNA recoveries than did a sample volume of 2.5 mL when assessed at the same conditions for all other variables. In general, increasing the sample volume from 1 to 2.5 mL decreased recovery by 3.3×10^4^ GU L^-1^. The concentration of magnetic beads in the sample also greatly impacted results, as RNA recovery increased by 5.7×10^4^ GU L^-1^ as the concentration of magnetic beads increased from 15 to 100 µL mL^-1^ sample.

Using optimal method conditions, recoveries of GI-SCV-2 spiked in wastewater samples at 1.8×10^4^ and 1.8×10^6^ GU L^-1^ were 86.1 and 4.6%, respectively. The method limit of detection (MLOD) for this optimized method in pre-filtered wastewater was determined to be 4.6×10^4^ GU L^-1^ with a 95% degree of confidence. The detection of SARS-CoV-2 in wastewater samples collected from the sewersheds of Communities A and C demonstrated that with viral RNA concentrations above the MLOD, the magnetic bead-based extraction protocol allowed rapid identification of SARS-CoV-2 in wastewater without the need for sample pre-concentration or centrifugation.

The optimized method described in this work may be used as an essential tool for the wastewater surveillance of SARS-CoV-2. The method protocol is simple and transferable, thus offering advantageous application for laboratories or circumstances that may have limited resources.

## Data Availability

All applicable data available in manuscript text

## 6. ACKNOWLEDGEMENTS

This study was funded through support from an NSERC COVID-19 Alliance Grant [grant number ALLRP 554503-20], an NSERC Collaborative Research and Development Grant in partnership with Halifax Water [grant number CRDPJ 539387-19] and the NSERC/Halifax Water Industrial Research Chair program [grant number IRCPJ: 349838-16]. Wastewater was spiked at the National Microbiology Laboratory in Winnipeg with GI-SCV-2 and HCV 229E acquired from the University of Alberta through the Canadian Water Network COVID-19 Coalition. The authors thank LuminUltra Technologies for providing qPCR technology and for technical support that was led through Dr. Jordan Schmidt throughout the project. The authors would like to extend thanks to municipal staff in communities A, B, and C for sample collection. As well, the authors would like to extend thanks to researchers Paul Bjorndahl and Sebastian Munoz from the Centre for Water Resources Studies at Dalhousie University for providing technical reviews, data analysis, and data visualization during the study. Graphical abstract was adapted from “*Quantifying SARS-CoV-2 Virions in City Wastewater”*, by BioRender.com (2020); retrieved from https://app.biorender.com/templates.

